# Multimodal Image Dataset for AI-based Skin Cancer (MIDAS) Benchmarking

**DOI:** 10.1101/2024.06.27.24309562

**Authors:** Albert S. Chiou, Jesutofunmi A. Omiye, Haiwen Gui, Susan M. Swetter, Justin M. Ko, Brian Gastman, Joshua Arbesman, Zhuo Ran Cai, Olivier Gevaert, Chris Sadee, Veronica M. Rotemberg, Seung Seog Han, Philipp Tschandl, Meghan Dickman, Elizabeth Bailey, Gordon Bae, Philip Bailin, Jennifer Boldrick, Kiana Yekrang, Peter Caroline, Jackson Hanna, Nicholas R. Kurtansky, Jochen Weber, Niki A. See, Michelle Phung, Marianna Gallegos, Roxana Daneshjou, Roberto Novoa

## Abstract

With an estimated 3 billion people globally lacking access to dermatological care, technological solutions leveraging artificial intelligence (AI) have been proposed to improve access^1^. Diagnostic AI algorithms, however, require high-quality datasets to allow development and testing, particularly those that enable evaluation of both unimodal and multimodal approaches. Currently, the majority of dermatology AI algorithms are built and tested on proprietary, siloed data, often from a single site and with only a single image type (i.e., clinical or dermoscopic). To address this, we developed and released the Melanoma Research Alliance Multimodal Image Dataset for AI-based Skin Cancer (MIDAS) dataset, the largest publicly available, prospectively-recruited, paired dermoscopic- and clinical image-based dataset of biopsy-proven and dermatopathology-labeled skin lesions. We explored model performance on real-world cases using four previously published state-of-the-art (SOTA) models and compared model-to-clinician diagnostic performance. We also assessed algorithm performance using clinical photography taken at different distances from the lesion to assess its influence across diagnostic categories.

We prospectively enrolled 796 patients through an IRB-approved protocol with informed consent representing 1290 unique lesions and 3830 total images (including dermoscopic and clinical images taken at 15-cm and 30-cm distance). Images represented the diagnostic diversity of lesions seen in general dermatology, with malignant, benign, and inflammatory lesions that included melanocytic nevi (22%; n=234), invasive cutaneous melanomas (4%; n=46), and melanoma in situ (4%; n=47). When evaluating SOTA models using the MIDAS dataset, we observed performance reduction across all models compared to their previously published performance metrics, indicating challenges to generalizability of current SOTA algorithms. As a comparative baseline, the dermatologists performing biopsies were 79% accurate with their top-1 diagnosis at differentiating a malignant from benign lesion. For malignant lesions, algorithms performed better on images acquired at 15-cm compared to 30-cm distance while dermoscopic images yielded higher sensitivity compared to clinical images.

Improving our understanding of the strengths and weaknesses of AI diagnostic algorithms is critical as these tools advance towards widespread clinical deployment. While many algorithms may report high performance metrics, caution should be taken due to the potential for overfitting to localized datasets. MIDAS’s robust, multimodal, and diverse dataset allows researchers to evaluate algorithms on our real-world images and better assess their generalizability.

## 1. Introduction

Skin disease causes significant morbidity and mortality, and access to dermatological care is of global concern. Artificial intelligence (AI) has been proposed as a technological solution to improve skin disease diagnosis and triage. However, to date, most algorithms do not reflect the workflow of dermatologists.

Dermatologists assess skin lesions using both macroscopic clinical examinations and specialized dermoscopy imaging, while integrating key demographic and medical history data known to influence malignancy risk ^2,3^. This multimodal approach may drive the improved performance of in-vivo assessment compared to dermoscopic photography. Despite this inherent multimodality, most dermatology artificial intelligence (AI) algorithms have been unimodal. For example, at the time of writing, prospective dermatology computer vision AI studies have focused exclusively on either clinical or dermoscopic images^4–7^.

While multimodal approaches to dermatological diagnostics are complex and require meticulous dataset curation and image acquisition, they have shown promise in enhancing performance and generalizability over unimodal methods^8^. Studies have demonstrated the efficacy of combining clinical and dermoscopic images to improve performance for both binary and multi-class classification tasks ^9,10^. Incorporating clinical meta-data as an adjunct to image data has also been repeatedly explored, with mixed results in improving model performance ^11,12^. Furthermore, these multimodal classifiers demonstrate better generalizability across datasets compared to their unimodal counterparts.

Although multimodal strategies show great potential in improving diagnostic accuracy for skin conditions, their adoption is significantly limited by the scarcity of rigorous external benchmark evaluations and limited access to multimodal datasets. The barriers to widespread development and release of transparent, well-annotated multimodal datasets include increased burden to traditional clinical workflows, the laborious nature of annotating multiple images, and previously documented issues arising from institutional privacy policies and incentives that favor siloed datasets ^13^. In addition, this lack of comprehensive validation poses an obstacle to the clinical utility of these algorithms. For instance, external validation of dermoscopic image-based algorithms in the International Skin Imaging Collaboration (ISIC) Grand Challenge revealed significant accuracy drops due to shifts in disease distributions, poor performance on conditions not represented in training, and routine clinical artifacts like hair or pen markings. Moreover, these algorithms often failed to recognize images beyond their training scope ^14^. Similarly, our group previously showed that state-of-the-art (SOTA) clinical algorithms demonstrate marked performance declines on an independent retrospective benchmark database, especially for patients with Fitzpatrick V-VI skin tones ^15^.

Prior multimodal work has been aimed at pigmented lesion classification tasks, with retrospective curation limiting its applicability ^16,17^ The absence of *prospective* multimodal datasets for external validation across a variety of diagnoses hampers the AI field’s ability to uncover and address algorithmic vulnerabilities or biases. Consequently, this limitation impedes the trust in the generalizability and clinical viability of dermatology-AI models.

To address this gap, we introduce the Melanoma Research Alliance Multimodal Image Dataset for AI-based Skin Cancer (MIDAS), the first publicly available, prospectively-recruited, systematically-paired dermoscopic and clinical image-based dataset across a range of skin-lesion diagnoses. This dual-center dataset encompasses a wide array of skin lesions and includes well-annotated, patient-level, clinical metadata. It aims to more accurately mirror real-world clinical scenarios than retrospectively curated datasets and is enhanced by extensive histopathologic confirmation to ensure data integrity. We also report an evaluation of four SOTA algorithms using the MIDAS dataset to assess their “out-of-the-box” performances. Additionally, we illustrate MIDAS’s significance in multimodal dermatology diagnostics, addressing clinical implementation questions like optimal image distance for different models and laying the foundation for future multimodal AI development using MIDAS.

## 2. Results

### 2.1 Demographics and dataset characteristics

796 total patients were prospectively enrolled in the study across two healthcare systems, with 734 from Stanford and 62 from Cleveland Clinic. Cleveland Clinic primarily participated as an additional expert pigmented lesion/melanoma center, while the Stanford sites included predominantly general dermatology clinics along with an expert pigmented lesion/melanoma clinic clinics. The population had a male predominance (64.9%), with a mean age of 60.9 years. The majority of patients represented clinician-assessed Fitzpatrick skin tones I/II, with 16.8% representing Fitzpatrick III-VI skin tones (**Table 1**).

**Table 1.** Demographic characteristics of the cohort. Patients were recruited from both general dermatology and expert pigmented lesion/melanoma clinics at Stanford. Cleveland Clinic also participated as an additional expert pigmented lesion/melanoma center.

| Characteristic | All<br>(N=796) | Stanford<br>(N=734) | Cleveland Clinic<br>(N=62) |
| --- | --- | --- | --- |
| **Age** |  |  |  |
| Mean (SD) | 60.9 (16.7) | 61.0 (16.8) | 60.4 (16.4) |
| **Sex** |  |  |  |
| Male | 517 (64.9%) | 477 (65.0%) | 40 (64.5%) |
| Female | 279 (35.1%) | 257 (35.0%) | 22 (35.5%) |
| **Race** |  |  |  |
| White | 657 | 596 (81.2%) | 61 (98.4%) |
| Asian | 57 | 56 (7.6%) | 1 (1.6%) |
| African American | 7 | 7 (1.0%) | 0 |
| American Indian/Alaskan Native | 2 | 2 (0.3%) | 0 |
| Other | 43 | 43 (5.8%) | 0 |
| Unknown/decline to state | 30 | 30 (4.2%) | 0 |
| **Ethnicity** |  |  |  |
| Hispanic | 38 | 38 (5.2%) | 0 |
| Non-Hispanic | 730 | 670 (91.1%) | 60 (96.8%) |
| Unknown/decline to state | 28 | 26 (3.7%) | 2 (3.2%) |
| **Fitzpatrick Skin Type** |  |  |  |
| I | 43 (5.4%) | 32 (4.4%) | 11 (17.7%) |
| II | 619 (77.8%) | 570 (77.7%) | 49 (79.0%) |
| III | 78 (9.8%) | 77 (10.5%) | 1 (1.6%) |
| IV | 42 (5.3%) | 41 (5.6%) | 1 (1.6%) |
| V | 5 (0.6%) | 5 (0.7%) | 0 |
| VI | 2 (0.3%) | 2 (0.3%) | 0 |
| Unknown | 7 (0.9%) | 7 (1.0%) | 0 |
| **History of Melanoma** |  |  |  |
| Yes | 158 (19.8%) | 123 (16.7%) | 35 (56.5%) |
| No | 633 (79.5%) | 606 (82.6%) | 27 (43.5%) |
| Unknown | 5 (0.6%) | 5 (0.7%) | 0 |

Among all patients, we enrolled a total of 1290 unique lesions. This resulted in a total of 3830 images, which included 1187 dermoscopy images, 1198 clinical images taken from a distance of 15-cm, 1185 clinical images taken from 30-cm, and 260 virtual images from photos submitted by patients or their primary care physicians via the patient portal. Among these, 244 represented control lesions that were not biopsied, while 1046 (81.1%) were biopsied with histopathologic confirmation; the latter were subsequently used in the primary analysis for evaluation of model performance. We employed a total of 3162 images for primary analysis of model performances: 1048 dermoscopy images, 1056 clinical images taken from 15-cm away, 1045 clinical images taken from 30-cm away, and 13 from virtual encounters where lesions were ultimately biopsied.

The most common final histopathology-confirmed diagnosis was benign melanocytic nevus (22.4%), followed closely by basal cell carcinoma (20.3%) **(Table 2)**. 135 total surgically eligible melanocytic lesions were diagnosed, including 46 (4.4%) invasive melanomas, 47 (4.5%) melanomas-in-situ, and 42 (4.0%) other surgically-eligible melanocytic diagnoses, such as severely dysplastic nevi and melanocytomas. In addition to melanocytic lesions, the dataset represents a relatively broad distribution of non-melanocytic lesions representing benign, pre-malignant, and malignant diagnoses commonly encountered in clinical practice, such as seborrheic keratoses (8.0%), actinic keratoses (6.0%), fibrous papules (0.6%), dermatofibromas (1.5%), and vascular lesions (1.0%).

**Table 2.** Biopsied lesion characteristics. These diagnoses were biopsy-proven and interpreted by 3 board-certified dermatopathologists at Stanford and 6 dermatopathologists at Cleveland Clinic. A dermatopathology consensus conference reviewed any diagnosis of severely dysplastic melanocytic nevus or worse at Stanford, and any diagnosis of invasive melanomas at CCF.

| Diagnoses | All<br>(N=1046) | Stanford<br>(N=964) | Cleveland Clinic<br>(N=82) |
| --- | --- | --- | --- |
| Actinic keratosis | 63 (6.0%) | 63 (6.5%) | 0 |
| Dermatofibroma | 16 (1.5%) | 16 (1.7%) | 0 |
| Fibrous papule | 6 (0.6%) | 6 (0.6%) | 0 |
| Hemangioma | 11 (1.0%) | 10 (1.0%) | 1 (1.2%) |
| Melanocytic nevus* | 234 (22.4%) | 193 (20.0%) | 41 (50.0%) |
| Seborrheic keratosis | 83 (8.0%) | 81 (8.4%) | 2 (2.4%) |
| Basal cell carcinoma | 212 (20.3%) | 202 (21.0%) | 10 (12.2%) |
| Invasive melanoma | 46 (4.4%) | 38 (3.9%) | 8 (9.8%) |
| Melanoma-in-situ | 47 (4.5%) | 39 (4.0%) | 8 (9.8%) |
| Other Melanocytic lesions, surgically eligible | 42 (4.0%) | 38 (3.9%) | 4 (4.8%) |
| Severely dysplastic nevus | 37 | 34 | 3 |
| Melanocytomas and intermediate melanocytic tumors^ | 5 | 4 | 1 |
| Squamous cell carcinoma | 71 (6.8%) | 68 (7.1%) | 3 (3.7%) |
| Squamous cell carcinoma in-situ | 53 (5.1%) | 53 (5.5%) | 0 |
| Other (benign) | 141 (13.5%) | 139 (14.4%) | 2 (2.4%) |
| Other (malignant) | 6 (0.6%) | 5 (0.5%) | 1 (1.2%) |
| Other (non-neoplastic, inflammatory, infectious) | 15 (1.4%) | 13 (1.3%) | 2 (2.4%) |

Before releasing the dataset, we reviewed all images and performed additional preprocessing, which included cropping and further de-identification. Currently, all consented Stanford images are publicly released at [https://stanfordaimi.azurewebsites.net/datasets/f4c2020f-801a-42dd-a477-a1a8357ef2a5]

### 2.2 Clinician impressions

For all biopsied lesions at Stanford, the initial treating dermatologist submitted their top 5 diagnoses based on their rank-ordered clinical impression prior to obtaining the biopsy results. We analyzed top 1 and top 3 diagnoses. Using top-1 diagnosis alone, clinicians were overall 79.3% accurate at differentiating a malignant from a benign lesion, with sensitivity and specificity of 78.4% and 80.0%, respectively **(Figure 1)**. This increased to 91.3% accuracy when evaluating top-3 benign vs. malignant diagnosis **(Supplementary Table 1)**. This dataset aims to be relevant for triage tasks and biopsy decision scenarios outside of an expert pigmented lesion clinic, where ruling in a concerning lesion takes primacy over ruling out a non-concerning lesion. Thus, we defined accuracy for clinicians around identifying the possibility of malignancy rather than providing the exact diagnosis.

**Figure 1.**
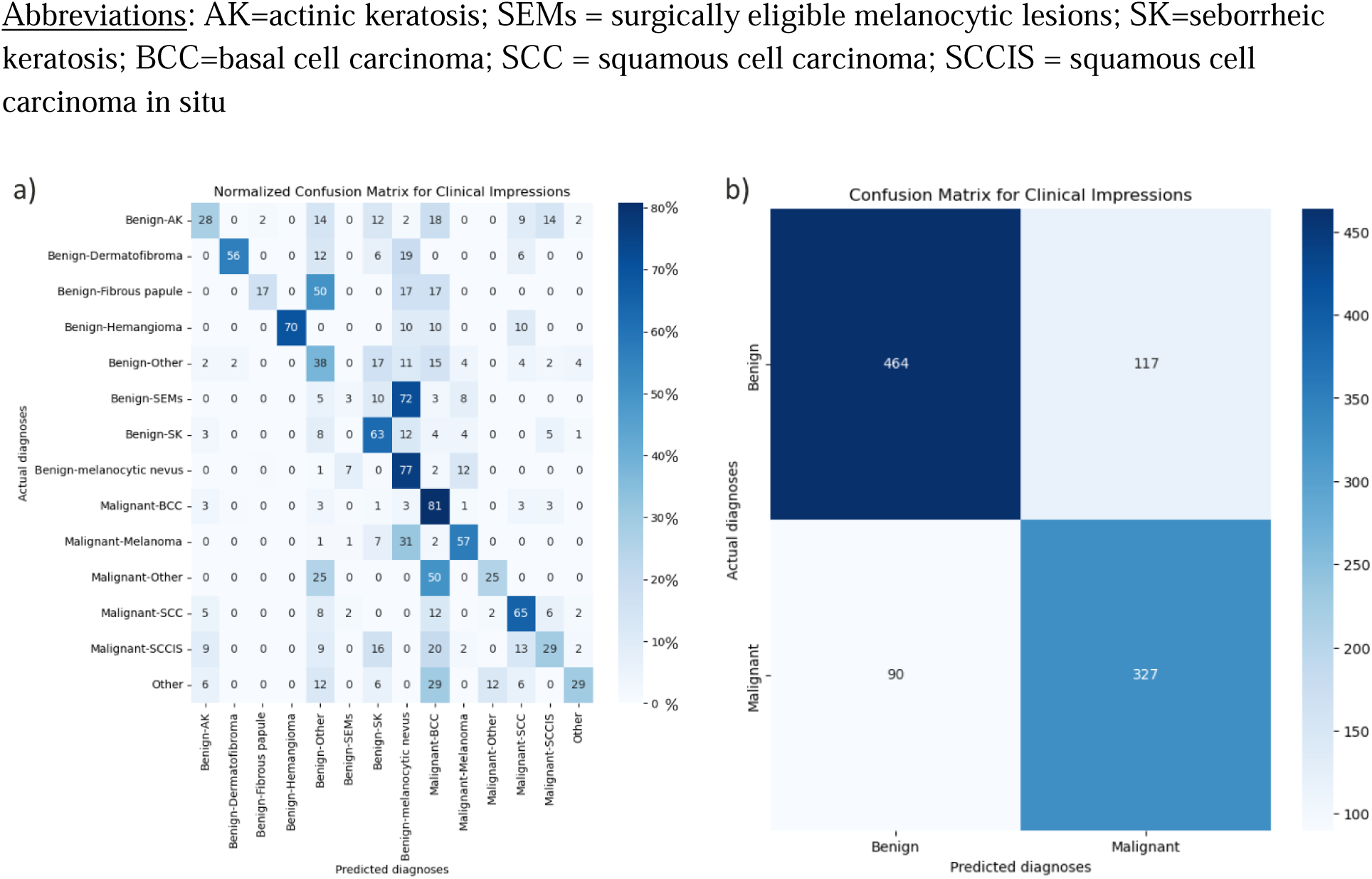
Confusion matrix for dermatologists’ top-1 diagnosis. a) Confusion matrix is normalized by actual diagnosis to allow for better visualization of clinician performance. b) Absolute numbers are provided for this confusion matrix. Benign lesions include: AK, dermatofibroma, fibrous papule, hemangioma, SEMs, SK, melanocytic nevus. Malignant lesions include: BCC, melanoma, melanoma-in-situ, SCC, SCCIS.

For the identification of melanomas (including invasive melanoma and melanoma-in-situ) for top-1 diagnosis, clinicians were 57% sensitive and 95% specific. When accounting for top-3 diagnoses, this increased to 94% sensitivity and 98% specificity. For top-3 diagnosis, clinicians were accurate if they mentioned the correct diagnosis in any of their top-3 differentials. We observed that most clinicians included both diagnoses of benign melanocytic neoplasms and malignant melanomas in their differentials for melanocytic lesions, making top-3 diagnosis less informative than top-1 diagnosis. Performance was high for the most common non-melanoma skin cancers, i.e., basal cell and squamous cell (keratinocyte) carcinomas. For basal cell carcinomas, using top-1 diagnosis, clinicians were 81% sensitive and 91% specific, while for invasive squamous cell carcinomas, clinicians were 65% sensitive and 97% specific using their top-1 diagnosis.

### 2.3 Benchmark Model Performance

Our evaluation revealed a performance decline for all models compared to their initial metrics, although the extent and dimensions of the decline varied significantly according to the model (**Figure 2**, **Table 3**).

**Figure 2.**
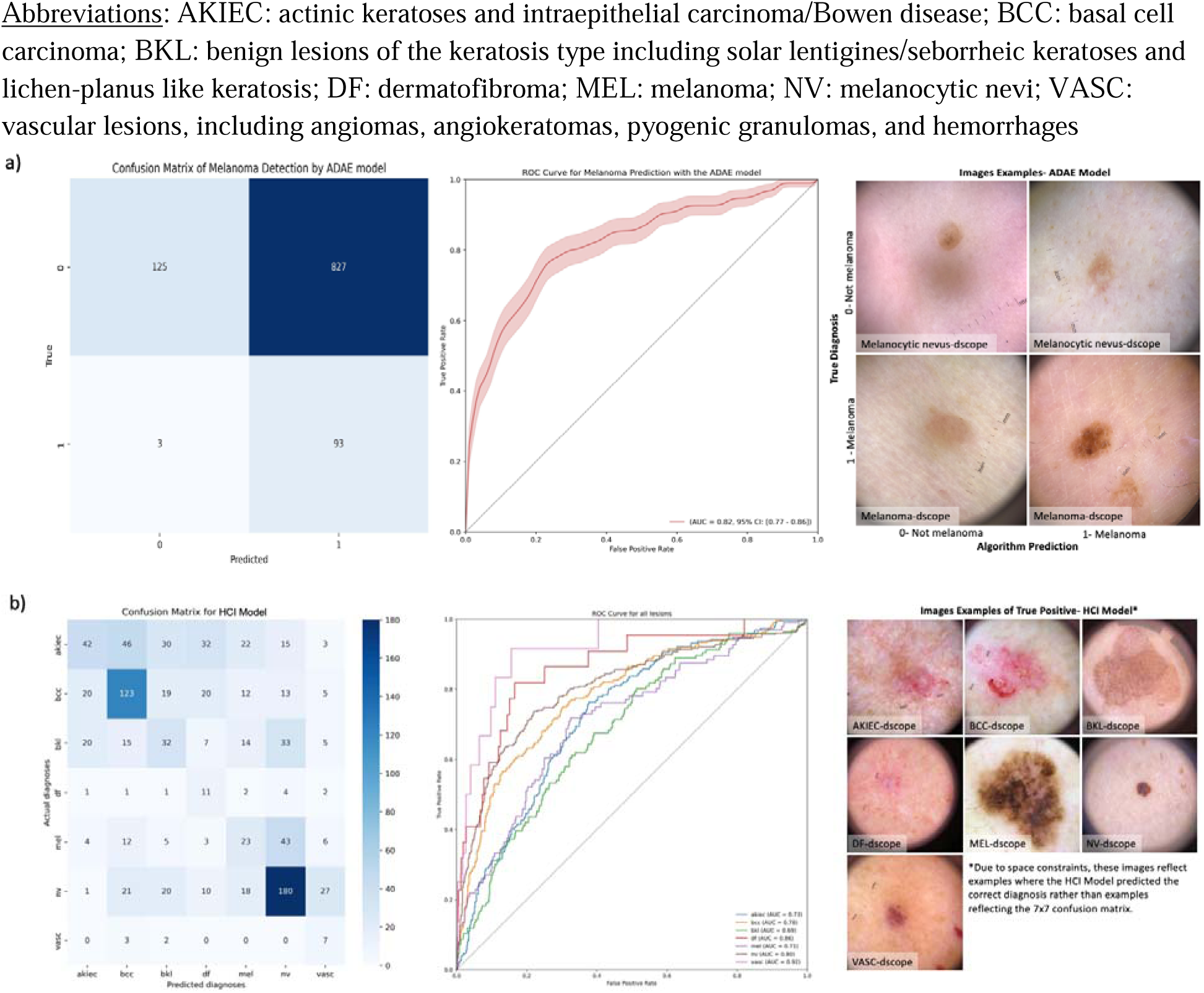
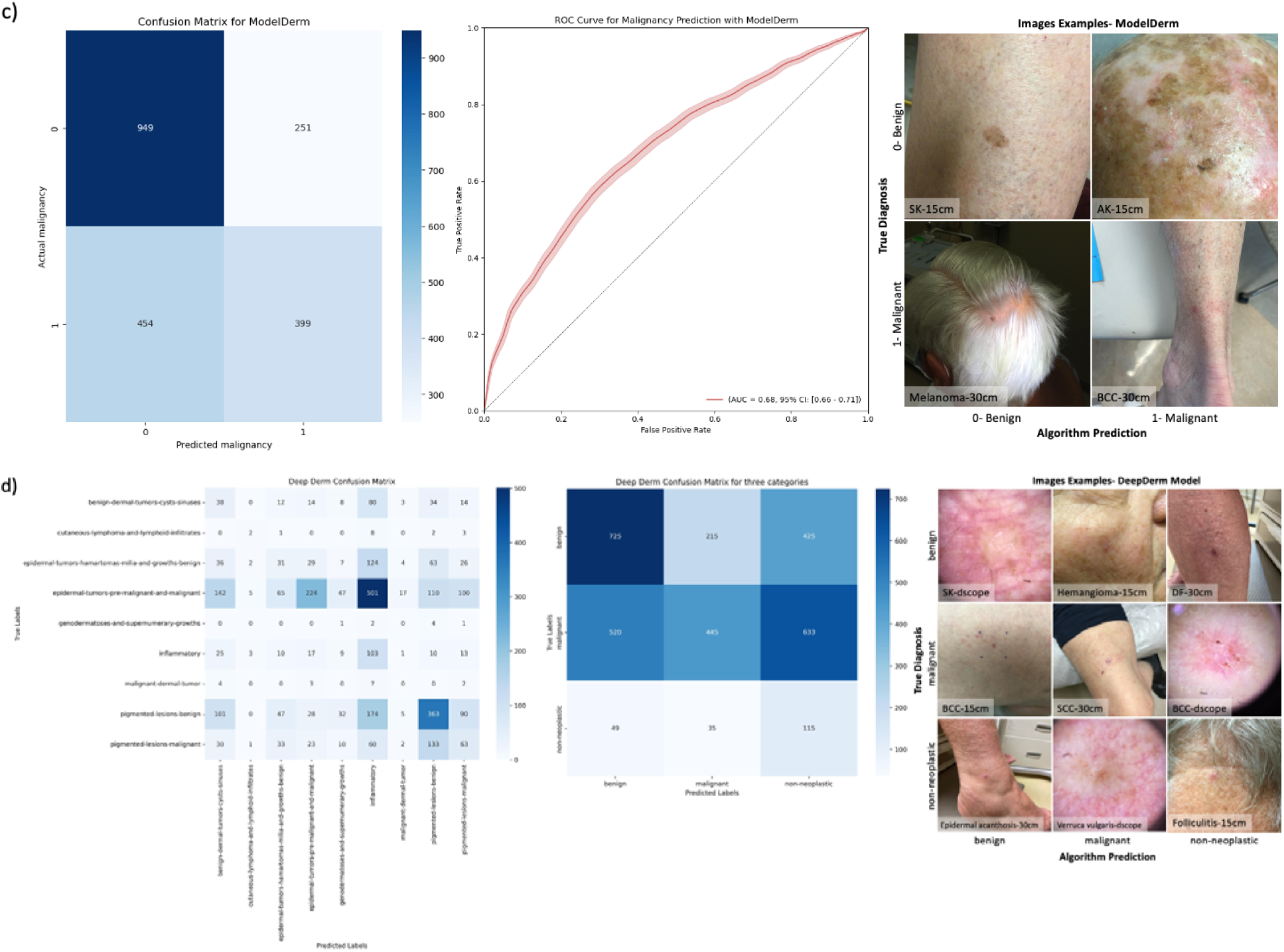
Model performance results-ROC curve, confusion matrix, and image examples. a) The evaluation metrics of the ADAE model, including the confusion matrix and ROC curve, alongside a random sample of the images that were correctly and incorrectly predicted are shown (far right). b) The performance of the HCI model is shown using the confusion matrix and multiclass ROC curve. The AUC for the ‘vasc’ category is the highest at 0.94. Dermoscopic images of correctly predicted lesions are also shown in the far-right for all the seven disease categories. c) ModelDerm’s performance is equally shown and samples of true positive (TP), true negative (TN), false negative (FN), and false positive (FP) clinical images are displayed. d) This shows the results of the DeepDerm model in the form of nine-label and three-label confusion matrices. Random image samples that correspond to correct and incorrect predictions are also displayed in a way that correlates with the three-label matrix.

**Table 3.** Model Performance. This depicts the performance of 4 state of the art models, which had varied inputs and use cases. We calculated the accuracy, sensitivity, specificity, precision and F1-score for each model.

| Model | Image Distance | Use Case | Class | Accuracy | Sensitivity | Specificity | Precision | F1 | AUROC [95% CI] |
| --- | --- | --- | --- | --- | --- | --- | --- | --- | --- |
| ADAE | Dscope | Melanoma classification | melanoma | 0.21 | 0.97 | 0.13 | 0.10 | 0.18 | 0.82 [0.77-0.8] |
| HCI | Dscope | Multi-class classification | akiec | 0.79 | 0.22 | 0.94 | 0.48 | 0.30 | 0.73 [0.69-0.7] |
|  |  |  | bcc | 0.80 | 0.58 | 0.86 | 0.56 | 0.57 | 0.78 [0.75-0.8] |
|  |  |  | bkl | 0.82 | 0.25 | 0.90 | 0.29 | 0.27 | 0.69 [0.64-0.7] |
|  |  |  | df | 0.91 | 0.50 | 0.90 | 0.29 | 0.27 | 0.86 [0.77-0.9] |
|  |  |  | mel | 0.85 | 0.24 | 0.92 | 0.25 | 0.25 | 0.71 [0.66-0.7] |
|  |  |  | nv | 0.79 | 0.65 | 0.84 | 0.63 | 0.64 | 0.80 [0.77-0.8] |
|  |  |  | vasc | 0.94 | 0.58 | 0.95 | 0.13 | 0.21 | 0.92 [0.84-0.9] |
| ModelDerm | 15cm, 30cm | Malignancy classification | malignant | 0.66 | 0.47 | 0.79 | 0.61 | 0.53 | 0.68 [0.66-0.7] |
| DeepDerm | Dscope, 15cm, 30cm | Three-category disease classification | benign | 0.62 | 0.53 | 0.68 | 0.56 | 0.55 | - |
|  |  |  | malignant | 0.56 | 0.28 | 0.84 | 0.64 | 0.39 | - |
|  |  |  | non-neoplastic | 0.64 | 0.58 | 0.64 | 0.10 | 0.17 | - |

The **ADAE model**, previously validated for melanoma diagnosis ^5^, showed a slight AUC decrease to 0.82 (from 0.857), with high sensitivity of 96.9% (95% CI: 91.2–98.9%) that compared favorably with the original model sensitivity of 96.8% (95% CI:91.1-98.9%). However, it showed significantly lower specificity of 13.1% (95% CI: 11.1–15.4%) and accuracy of 21% (95% CI: 18.5-23.4%) on the MIDAS dataset **(Table 3)**, primarily due to increased false positives. Focusing only on MIDAS’s melanocytic subset, ADAE’s sensitivity remained high, with specificity and accuracy slightly improved to 17.0% (95% CI: 13.0–21.8%), and 37.5% (95% CI: 32.8–42.6%) respectively.

Across all disease categories, the **HCI model** achieved sensitivity of 43.3% (95% CI: 29.5-55.7%) and specificity of 90.4% (95% CI: 87.8-92.9%), with overall accuracy of 84.2% (95% CI: 80.1-88.9%), which was similar to the original results reported by Tschandl et al (80.3%). For malignant lesions, the model’s sensitivity was 58.2% (95% CI: 56.9-65.2%), and specificity was 77.5% (95% CI: 74.1-81.9%), reflecting a drop in performance compared to its original study of 58.7% sensitivity and 93.3% specificity^19^.

**ModelDerm’s** performance on MIDAS’s distance of 15-cm and 30-cm clinical images revealed an AUC of 0.68 (95% CI: 0.66-0.71), which was notably lower than its retrospective validation results of 0.86 (95% CI: 0.85-0.88). The model’s sensitivity and specificity for malignancy detection were 46.8% and 79.0%, respectively, indicating a notable variation from the original 62.7% (95% CI 59.9–65.1) sensitivity and 90.0% (95% CI 89.4–90.6) specificity ^24^.

**DeepDerm’s** mean accuracy for the three-way classification task dropped to 43.3% (**Table 3**), which was a significant decrease from the initially reported 72.1% ^23^. Its sensitivity for malignant diagnoses was low at 28.0%, with a specificity of 84.1%. Misclassification patterns revealed that malignant lesions were most commonly misclassified as non-neoplastic (inflammatory), followed by benign. We also explored performance discrepancies across the different hospital sites (**Supplementary Table 2)**.

#### 2.3.1 Poor performers

Within this dataset, 27 lesions were identified as the most challenging cases, since all four models incorrectly classified every tested image at top-1 diagnosis. Clinicians demonstrated a 51.9% accuracy as top-1 diagnosis within this challenging subset of lesions. Basal cell carcinomas and squamous cell carcinoma in situ comprised the majority (20/27) of these incorrectly classified images, but there was one melanoma within this dataset that was incorrectly predicted to represent a nevus by the clinicians as a top-1 diagnosis.

#### 2.3.2 Optimizing Image Acquisition Using Paired Camera Distances

The optimal camera distance of image acquisition for algorithm performance was evaluated using both Deepderm and Modelderm, because both utilized clinical images either exclusively as one of the primary modalities in their training. When looking at the effects of camera distance on model performance, accuracy, and sensitivity improved for benign and malignant lesions with images acquired at 15-cm photography as compared to 30-cm photography across both models **(Table 4)**. This was especially notable for DeepDerm in the classification of malignant lesions, with an improvement in sensitivity from 0.17 to 0.30. Overall accuracy for non-neoplastic disease classification by DeepDerm improved at 15-cm photography due to improved specificity but at a significant cost to sensitivity.

**Table 4.** Model subanalysis: Image distance. We explored the impact of image distance-dermoscopy, 15cm and 30cm on ModelDerm and DeepDerm performance. ModelDerm was only trained on clinical images; DeepDerm was trained on both dermoscopy images and clinical images.

| Model | Class | Image Distance | Accuracy | Sensitivity | Specificity | Precision | F1-Score | AUROC [95% CI] |
| --- | --- | --- | --- | --- | --- | --- | --- | --- |
| ModelDerm | malignant | dermoscope | 0.62 | 0.39 | 0.78 | 0.56 | 0.46 | 0.65 [0.62-0.69] |
|  |  | **15cm** | **0.67** | **0.49** | **0.79** | **0.63** | **0.55** | **0.70 [0.66-0.73]** |
|  |  | 30cm | 0.65 | 0.44 | 0.79 | 0.59 | 0.51 | 0.67 [0.64-0.71] |
| DeepDerm | benign | **dermoscope** | **0.63** | **0.62** | **0.64** | **0.57** | **0.59** | - |
|  |  | 15cm | 0.62 | 0.57 | 0.66 | 0.56 | 0.57 | - |
|  |  | 30cm | 0.60 | 0.41 | 0.75 | 0.55 | 0.47 | - |
|  | malignant | dermoscope | 0.56 | 0.37 | 0.76 | 0.62 | 0.46 | - |
|  |  | 15cm | 0.57 | 0.30 | 0.84 | 0.66 | 0.41 | - |
|  |  | 30cm | 0.54 | 0.17 | 0.92 | 0.68 | 0.27 | - |
|  | non-neoplastic | **dermoscope** | **0.78** | **0.56** | **0.79** | **0.14** | **0.23** | - |
|  |  | 15cm | 0.66 | 0.45 | 0.68 | 0.09 | 0.14 | - |
|  |  | 30cm | 0.48 | 0.74 | 0.46 | 0.09 | 0.15 | - |

#### 2.3.3 Image modality: Dermoscopy vs. clinical images

Of the four algorithms, DeepDerm was the only one that incorporated both dermoscopy images and clinical images in its training, and it thus provided the best vehicle to evaluate the comparative performance of dermoscopy and clinical images with a single algorithm ^23^. As expected, we confirmed that ModelDerm performed worse on dermoscopy images compared to clinical images, since it was trained solely on clinical images **(Table 4**). In contrast, DeepDerm in general demonstrated its best performance on dermoscopy images, with notable improvement in its identification of non-neoplastic lesions. For the classification of malignant lesions, DeepDerm showed improvement in sensitivity from 0.30 to 0.37 moving from 15-cm clinical photography to dermoscopic images, with minimal impact on overall accuracy, suggesting a potential slight benefit for dermoscopy over clinical images for triage-based tasks where a higher sensitivity is favorable, though at the cost of slightly reduced specificity.

#### 2.3.4 Clinical characteristics influencing performance

We explored the effects of demographics and other clinical information on model performance, including patient natal sex, age, prior history of melanoma, Fitzpatrick skin type, and lesion size **(Supplementary Table 3)**. We noted that ADAE performed better in patients with a prior history of melanoma (accuracy 28% vs 19% in those with no prior history), likely due to a higher prevalence of subsequent primary melanomas in patients with a history of melanoma. DeepDerm also performed better in lesions where patients had a prior history of melanoma (accuracy 73% vs 62% in patients with no prior history). We also found that ModelDerm performed better in lesions with Fitzpatrick skin type 3-6 (accuracy of 78% vs 59% in Fitzpatrick skin types 1-2). This improved performance of patients with darker skin types was maintained but with a smaller magnitude difference when we compared F1-scores, which are better evaluation metrics to compare subgroups with class imbalances (F1-score of 0.57 in Fitzpatrick 3-6 vs 0.53 in Fitzpatrick 1-2).

#### 2.3.5 Ensemble models with multiple modalities

As an exploratory analysis, we investigated the performance of multimodal evaluation utilizing both dermoscopic and clinical image input through ensembling of the outputs of two of the previously published SOTA algorithms (see Methods 4.4.3 Proof-of-Concept Ensemble Models). As shown in **Table 5**, ensemble models A and B demonstrated increased accuracy at a cost of decreased sensitivity compared to the baseline models, ADAE and ModelDerm. Specifically, Ensemble Model A and B had accuracies of 75% compared to ADAE’s baseline of 37% and ModelDerm’s 74%. Notably, our logistic regression meta-model showed improved performance across multiple evaluation metrics with an AUC of 0.86 (95% CI: 0.76-0.94), accuracy of 82%, and specificity of 0.96.

**Table 5.**
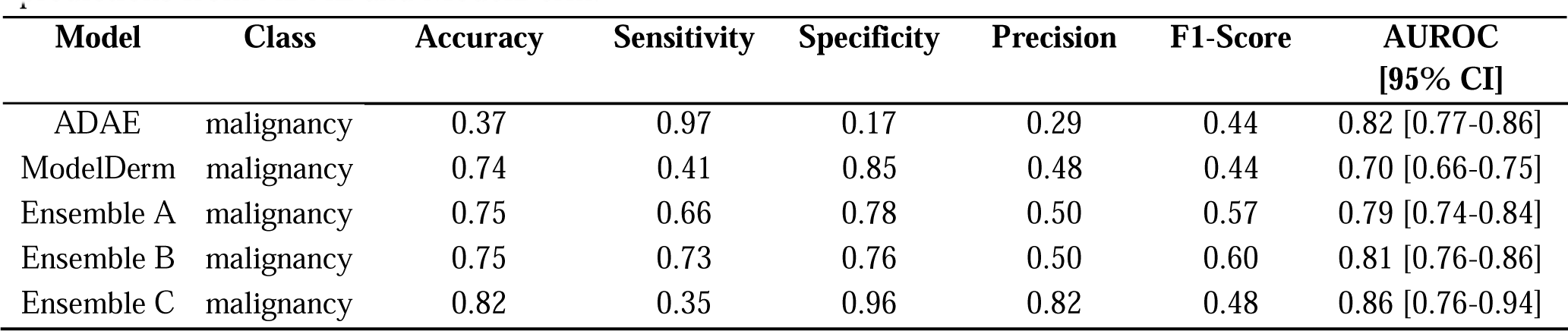
Ensemble Model Performance on Melanocytic Lesions. We explored model performance and ensemble models only on melanocytic lesions (melanocytic nevus, invasive melanoma, melanoma-in-situ, and surgically eligible melanocytic lesions). Ensemble Model A had weights of ⅓ ADAE dermoscopy + ⅓ ModelDerm 15cm clinical image + ⅓ ModelDerm 30cm clinical image. Ensemble Model B had weights of ½ ADAE dermoscopy + ¼ ModelDerm 15cm clinical image + ¼ ModelDerm 30cm clinical image. Ensemble Model C is a meta-model that employs logistic regression to learn the weights of the predictions from ADAE and ModelDerm.

## 3. Discussion

The translation of AI diagnostic tools into clinical practice, particularly in dermatology, presents a uniquely challenging and instructive paradigm due to the broad distribution of diagnoses and range of influencing factors such as skin tone, presence of artifacts, and variability in image acquisition. As these tools advance toward wider clinical deployment, the task of benchmarking and comprehensively understanding the strengths and vulnerabilities of SOTA diagnostic algorithms is increasingly paramount. The recent landmark U.S. “Executive Order on the Safe, Secure, and Trustworthy Development and Use of Artificial Intelligence” emphasized the importance of assuring the quality of medical AI^25^. While the debate continues over the most valuable form of validation—be it ‘gold standard’ external validation or local validation at the individual facility level ^26^, it is clear that independent, pre-deployment assessments are an important way to evaluate the generalizability and reliability of these algorithms.

Studies have shown that even within dermatologic practices, changing the intended clinical setting and distribution of diagnoses can significantly degrade algorithm performance, with the most striking performance reductions seen in more real-world, prospective contexts ^14,15,20^. While these studies represent progress towards external validation for unimodal lesion classifiers, the lack of well-annotated, transparent multimodal datasets is a major impediment to the ultimate development of multimodal diagnostic tools. In this context, the MIDAS dataset enables the utility of multiple modalities, along with meta-data inputs for both pigmented and non-pigmented skin lesions, and it covers a breadth of benign, pre-malignant, and malignant lesion types.

A key strength of our dataset is that it offers applicability in the evaluation of algorithms intended for diagnostic settings where a broader distribution of lesions is expected. These include general dermatology clinics, teledermatology-based practices, and primary care clinics. The prospective nature of MIDAS invariably included enrollment of lesions with secondary changes such as hemorrhagic crusts, excoriations, or edema from inflamed nevi or irritated seborrheic keratoses which have been shown to reduce algorithm performance ^14^. This inclusion supports the evaluation of algorithms on these common, real-world image artifacts inherent to the diseases and patients. A limitation of the dataset is the lower representation of patients with Fitzpatrick IV-VI skin types. This likely resulted from both the lower incidence of skin cancer in people of color and potentially disparate access to care at academic medical centers. We anticipate a solution to this important issue is increasing the availability of transparent, publicly available datasets where skin cancer cases in patients of color can be increasingly aggregated, and we believe that reporting MIDAS’s skin tone distribution and its public release can contribute to this effort. There are additional efforts in the community to increase knowledge about diagnostic imaging in skin of color^27^. In this respect, ModelDerm’s outperformance of the other algorithms on patients with darker Fitzpatrick types may be due in part to its training data, which was generated in large part in South Korea, thus highlighting the potential impact of the demographics of patients represented within algorithms’ training on their ultimate performance across diverse populations.

In our benchmarking experiments using multiple SOTA models utilizing a variety of inputs, we observed a decline in key performance measures, particularly sensitivity for malignant diagnoses and overall accuracy, across three of the four algorithms when deployed ’out of the box’, without threshold adjustments or fine-tuning. This significant drop in model performance underscores the necessity for comprehensive pre-deployment analysis and local optimization prior to the clinical deployment of dermatologic classifiers. Moreover, it demonstrates how imperative ongoing performance monitoring is to ensure the model remains within the local control parameters, and highlights a potential regulatory challenge. Establishing regulatory frameworks that permit and govern such continuous monitoring will be crucial. From the algorithm development perspective, with the exception of ADAE, the algorithms were trained predominantly on data from a select number of institutions (typically one to three), hence broadening the institutional and geographic basis of future diagnostic algorithms may improve generalizability. One particularly striking pattern of errors revealed through this dataset was in the DeepDerm algorithm, where epidermal tumors were consistently miss-classified as non-neoplastic (inflammatory) lesions, suggesting the need for further fine-tuning to the task at hand. Finally, the development of MIDAS provides an instructive case in how performance can be negatively impacted when classifiers that were trained primarily on pigmented lesion classification are challenged with a broader disease distribution. This is of particular value for evaluating algorithms intended for practitioners assessing a wider variety of lesional diagnoses.

Of note, both of the exclusively dermoscopic algorithms, HCI and ADAE, maintained reasonably robust AUCs in their respective classification tasks, implying significant potential benefit from pre-deployment optimization. ADAE, uniquely among the deployed algorithms, maintained a very high sensitivity for melanoma due to its design around a pre-specified 95% sensitivity threshold. However, with our broader dataset, it demonstrated a markedly reduced specificity of 13.1% (95% CI: 11.1–15.4%) which slightly increased to 17.0% (95% CI: 13.0–21.8%) in the subanalysis limited to melanocytic lesions. These results highlight the importance of understanding and adhering to the task for which an algorithm was originally trained, as ADAE was intended for use in cases where melanoma was in the differential diagnosis and thus provides post-clinician decision support to minimize unnecessary biopsies/false positives rather than ‘triaging in’ potentially concerning lesions. Challenging the algorithm with non-melanocytic cases and other out-of-distribution clinical scenarios likely contributed to the observed change in performance characteristics.

Our evaluation of ensemble approaches on MIDAS also yielded promising results, underscoring the dataset’s utility in assessing multimodal strategies. These findings stress the importance of not only pre-deployment validation, but also post-deployment surveillance and regulatory aspects of maintaining algorithmic efficacy and safety in real-world intended settings. Our study is not without limitations, however, including restricted access to some models, which constrained our ability to explore fusion approaches and fully leverage the multimodal dataset for evaluation. In conclusion, this novel, publicly available MIDAS dataset can be utilized as a valuable tool in dermatology-AI, offering a rigorous benchmark for refining multimodal diagnostic models and addressing the complexities of real-world clinical applications.

## 4. Methods

### 4.1 Patient enrollment and image acquisition

All research was approved by the Institutional Review Board at Stanford University under IRB#36050, along with the Cleveland Clinic Foundation under IRB#20-666, and adhered to the Helsinki Declaration. Patients presenting to the dermatology clinics of participating dermatologists at Stanford Medicine or Cleveland Clinic Foundation between August 18, 2020, and April 17, 2023, were eligible for the study if 1) they had at least one solitary skin lesion of concern identified where a skin biopsy was deemed medically necessary by the dermatologist investigator or 2) patients were directed to in-clinic evaluation for a lesion that was previously identified as concerning through a teledermatology encounter or dermatologist review of a patient photo submitted through the electronic patient messaging portal. Patients underwent written informed consent with either the physician or research coordinator, after which both clinical and dermoscopic digital photography were obtained of any eligible skin lesions. Each lesion underwent standardized photography with a contemporary model iPhone or iPad device (iPhone SE to iPhone 12 Pro and iPod touch to iPad mini) without flash photography at 15-cm and 30-cm distances, along with digital dermatoscope photography. For each lesion, clinical information about the patient was obtained and recorded including sex assigned at birth, age, Fitzpatrick skin type, personal history of melanoma, anatomic location, and the lesion’s length and width. Investigators had the discretion to identify additional control lesions that clinically appeared benign on a corresponding contralateral body site that were similarly enrolled for digital photography as an un-biopsied control lesion to include in the dataset, though primary analysis was restricted to biopsied lesions.

Nine Stanford dermatologists, two Cleveland Clinic dermatologists, and one Cleveland Clinic surgical oncologist recruited patients primarily from adult general medical dermatology clinics (Stanford) along with a subset (12.6%) from specialized pigmented lesion and melanoma clinics (Stanford and Cleveland). At the time of first enrollment, the four Stanford dermatologists at the specialized pigmented lesion and melanoma clinic had an average of 15.7 years of post-residency experience, while those in general medical dermatology clinics had an average of 3.9 years’ experience. Cleveland Clinic dermatologists/surgical oncologists had an average of 25.5 years of post-residency experience.

Dermatologists noted their top five ranked clinical impressions at the time of evaluation, along with their binary level of confidence (Yes/No) in their top impression.

### 4.2 Reference labeling

For any biopsied lesions, associated histopathologic final diagnoses were recorded and categorized into a previously described taxonomy (ZC, AC, RN). Biopsy results were interpreted by three board-certified dermatopathologists at Stanford and 6 dermatopathologists at Cleveland Clinic. A dermatopathology consensus conference reviewed any diagnosis of severely dysplastic melanocytic nevus or worse at Stanford, and any diagnosis of invasive melanomas at CCF. Melanocytic lesions were specifically grouped in the following manner: benign melanocytic nevi, melanomas (including melanoma in-situ and invasive melanoma), and surgically-eligible intermediate melanocytic tumors where complete excision is typically recommended at Stanford and Cleveland Clinic (including severely dysplastic melanocytic nevi and melanocytomas such as typical/atypical Spitz tumors, BAP-1-inactivated melanocytic tumors, deep penetrating nevi/tumors, and cellular blue nevi with atypia). For lesions deemed as controls or not biopsied, the top favored diagnosis by the original treating physician was chosen as the reference label. Cases were included in the dataset if a second reviewing independent board-certified dermatologist (ZC) agreed with the favored diagnosis based on a review of the associated images.

### 4.3 Data preprocessing and exploratory analysis

For all images collected, we focused on the subset of lesions that were biopsied with a pathology report. We cleaned the dataset by ensuring consistency across demographic labeling and correcting any missing data in the metadata. We manually parsed through all images in the dataset to ensure image quality. To better understand our dataset, we conducted exploratory analyses on patient demographics and lesion diagnostic breakdown. To maintain consistency across our analysis, for melanocytic lesions, we categorized invasive melanoma and melanoma-in-situ as malignant lesions. All other melanocytic lesions, including surgically-eligible melanocytic lesions, were categorized as benign lesions. In contrast, the SOTA algorithms (see section below for algorithm details) had different classifications of actinic keratosis; ADAE and ModelDerm categorized these lesions as benign, while the HCI model and DeepDerm classified these as malignant. In our analysis, we categorized actinic keratoses (AKs) as clinically benign, but for the HCI and DeepDerm models we kept the original classification and noted them as malignant. This difference in classification yields a unique perspective where the categorization of a lesion can be influenced by model selection - an academic task of differentiating between malignant and malignant-potential lesions, versus a triage-based task of identifying which lesions may need to be biopsied to rule out malignancies.

### 4.4 Evaluating models

Given our dataset’s prospectively-recruited and multimodal nature, we explored its potential use as a benchmark tool for external validation and comparative analysis of algorithms that leverage different elements of MIDAS’s image and clinical metadata. Specifically, we examined the performance of four previously published SOTA models, each employing distinct input modalities as described in subsequent sections, to understand their performance on this prospective data. Our analysis included two models based on dermoscopy images, one model predicated on clinical images, and one model designed to process either clinical or dermoscopic images.

To assess their ’out of the box’ performance, we adhered to the threshold values as stipulated by the original models. All four models were tested on the entire biopsy-proven analysis dataset. However, the evaluation of each model was tailored to the primary classification task for which it was initially designed, such as the binary classification of melanoma vs. non-melanoma, or malignant vs. benign lesions, and multi-class categorization.

#### 4.4.1 Dermoscopy-based Models

##### All Data are External (ADAE)

The ADAE algorithm is an open-source, non-commercial dermoscopic model that emerged as the top-ranking algorithm in the ISIC 2020 melanoma classification challenge ^5^. This ensemble model comprises 18 separate CNN-based models, which were trained on 5-folds totaling 90 models, each contributing to a collective probability class label. Notably, four model architectures incorporate metadata fields (age, sex, anatomic location, width, and height of images) to enhance prediction accuracy. Initially designed as a melanoma screening tool, the ADAE was thresholded to achieve high sensitivity (above 95%) for melanoma detection. We focused on a version of the model recently detailed by Marchetti et al.^5^, which demonstrated an AUC of 0.857 and sensitivity of 96.8% (95% CI: 91.1-98.9%) in a prospective validation study.

Our evaluation involved 1046 dermoscopic images from our dataset, examining the model’s proficiency in identifying melanoma, defined inclusively as invasive melanoma and melanoma-in-situ. Additionally, we performed a focused sub-analysis on melanocytic lesions to closely align with the ADAE model’s primary use case, where melanoma is considered a key differential diagnosis. Consistent with its primary training objective, a prediction by the modified ADAE was deemed accurate if the corresponding true label was either melanoma-in-situ or invasive melanoma, aligning with the model’s intended triage role.

##### HCI (Human–Computer Interaction) Model

The HCI model, developed by Tschandl et. al, is a ResNet34-based architecture that was fine-tuned for multi-class classification on the HAM10000 pigmented lesions dataset ^18^. The original model outputs probabilities for seven categories: actinic keratoses and intraepithelial carcinoma/Bowen disease (akiec), basal cell carcinoma (bcc), benign lesions of the keratosis type including solar lentigines/seborrheic keratoses and lichen-planus like keratosis (bkl), dermatofibroma (df), melanoma (mel), melanocytic nevi (nv) and vascular lesions, including angiomas, angiokeratomas, pyogenic granulomas, and hemorrhages (vasc)^19^. In this study, we evaluated the model on our prospectively-collected dermoscopic dataset.

Images of rare diseases not represented by the HCI model’s predefined categories were excluded to maintain category consistency. All pathologic diagnoses were mapped to the seven-category framework and discrepancies were resolved by two board-certified dermatologists (R.N. and R.D.). Following this mapping process, we ran the model in test mode on 935 dermoscopic images: nv: 276, bcc: 212, akiec: 190, bkl: 124, mel: 97, df: 22, vasc: 14. Both category-specific and overall performance metrics like AUC, sensitivity, and accuracy were calculated. Additionally, specific metrics for malignant cases were reported, providing a comprehensive overview of the model’s capabilities.

#### 4.4.2 Clinical Image and Clinical Image + Dermoscopy-based Models

##### ModelDerm

ModelDerm, an ensemble of ResNet variants that was trained on clinical images of skin lesions, can classify 184 skin diseases ^20,21^. Similar to ADAE and HCI models, we based our methodology on Han et al. ^20,21^, using the same thresholds as defined by their study. We used the 2019 build version of the model. The malignancy score was calculated as the sum of malignant outputs and 0.2[×[premalignant outputs as previously defined^22^. The algorithm consists of two modules: a lesion detector followed by the ResNet ensemble classifier. We excluded 3% (98/3162) of images where ModelDerm was unable to detect a lesion.

##### DeepDerm

The DeepDerm model, developed similarly to Esteva et al.’s algorithm, was trained on a comprehensive dataset including images from the ISIC Dermoscopic Archive, the Edinburgh Dermofit Library, and Stanford Medicine ^23^. Employing the ResNet50 architecture, the model was trained to classify skin conditions into a detailed 836-category taxonomy, which also simplifies into nine and three-category frameworks representing various dermatological conditions ^23^.

Herein, we aligned pathology reports from our prospectively collected dataset with the DeepDerm taxonomy, resolving discrepancies through a consensus approach among two board-certified dermatologists (R.N. and R.D.). The model’s performance, including category-specific and overall metrics, was evaluated on the dataset, alongside examining its performance across the two participating centers.

#### 4.4.3 Proof-of-Concept Ensemble Models

We further investigated the potential use of the MRA-MIDAS dataset for the development of multimodal ensemble approaches. We developed a proof-of-concept weight-based ensemble model using ADAE and ModelDerm, focusing on dermoscopy and clinical images. Due to the proprietary nature of the models, our ensemble approach was limited to the final output layers, employing two strategies: 1) a weighted average model and 2) a logistic regression-based meta-model. The weighted average model assigned: a) equal weights, distributing ⅓ each to ADAE dermoscopy, ModelDerm 15-cm and 30-cm predictions. b) higher weight to ADAE (½) with the remaining ½ equally divided between ModelDerm 15-cm and 30-cm predictions.

For the meta-model, we utilized a logistic regression algorithm to integrate predictions from ADAE and ModelDerm (15-cm and 30-cm). It was optimized via grid search over various regularization parameters, and we employed five-fold cross-validation for training. The evaluation metrics were based on a held-out test set. For consistent comparison in our ensemble models, our analyses were conducted on melanocytic lesions (melanocytic nevus, invasive melanoma, melanoma-in-situ, and surgically eligible melanocytic lesions).

#### 4.4.4 Statistical Analysis

Confidence intervals were calculated using the bootstrapping method and all analyses were performed using Python (version 3.8), and Scikit-learn (version 1.2).

## Acknowledgments

This material is the result of work supported with resources and the use of facilities at the Veterans Affairs Palo Alto Health Care System in Palo Alto, California. We would like to acknowledge Vijaytha Muralidharan, Susan Weber, and other team members who have helped support this work.

## SUPPLEMENTARY

**Supplementary Table 1.**
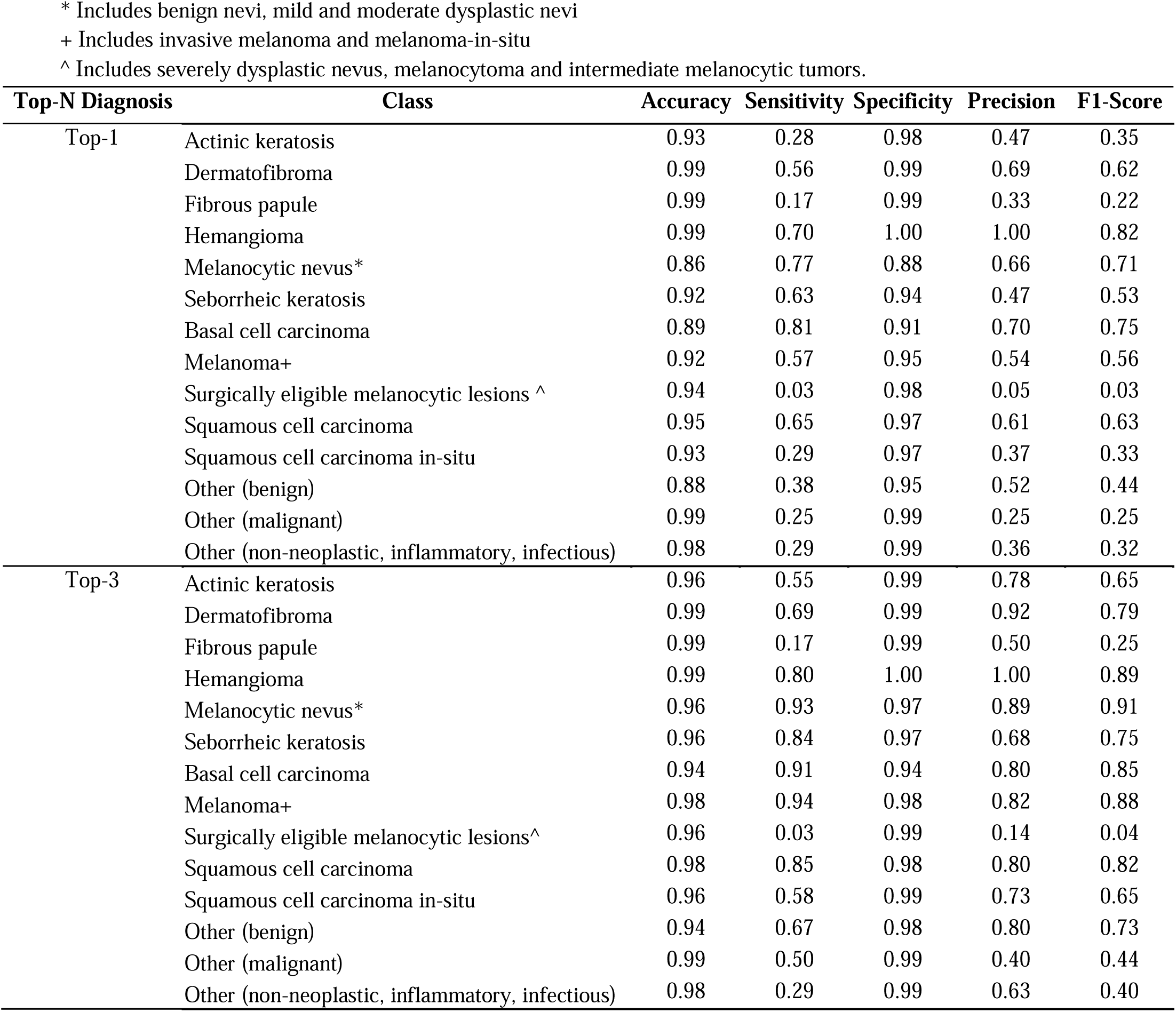
Clinician Impressions-Top1 and Top3.

| Top-N Diagnosis | Class | Accuracy | Sensitivity | Specificity | Precision | F1-Score |
| --- | --- | --- | --- | --- | --- | --- |
| Top-1 | Actinic keratosis | 0.93 | 0.28 | 0.98 | 0.47 | 0.35 |
|  | Dermatofibroma | 0.99 | 0.56 | 0.99 | 0.69 | 0.62 |
|  | Fibrous papule | 0.99 | 0.17 | 0.99 | 0.33 | 0.22 |
|  | Hemangioma | 0.99 | 0.70 | 1.00 | 1.00 | 0.82 |
|  | Melanocytic nevus* | 0.86 | 0.77 | 0.88 | 0.66 | 0.71 |
|  | Seborrheic keratosis | 0.92 | 0.63 | 0.94 | 0.47 | 0.53 |
|  | Basal cell carcinoma | 0.89 | 0.81 | 0.91 | 0.70 | 0.75 |
|  | Melanoma+ | 0.92 | 0.57 | 0.95 | 0.54 | 0.56 |
|  | Surgically eligible melanocytic lesions ^ | 0.94 | 0.03 | 0.98 | 0.05 | 0.03 |
|  | Squamous cell carcinoma | 0.95 | 0.65 | 0.97 | 0.61 | 0.63 |
|  | Squamous cell carcinoma in-situ | 0.93 | 0.29 | 0.97 | 0.37 | 0.33 |
|  | Other (benign) | 0.88 | 0.38 | 0.95 | 0.52 | 0.44 |
|  | Other (malignant) | 0.99 | 0.25 | 0.99 | 0.25 | 0.25 |
|  | Other (non-neoplastic, inflammatory, infectious) | 0.98 | 0.29 | 0.99 | 0.36 | 0.32 |
| Top-3 | Actinic keratosis | 0.96 | 0.55 | 0.99 | 0.78 | 0.65 |
|  | Dermatofibroma | 0.99 | 0.69 | 0.99 | 0.92 | 0.79 |
|  | Fibrous papule | 0.99 | 0.17 | 0.99 | 0.50 | 0.25 |
|  | Hemangioma | 0.99 | 0.80 | 1.00 | 1.00 | 0.89 |
|  | Melanocytic nevus* | 0.96 | 0.93 | 0.97 | 0.89 | 0.91 |
|  | Seborrheic keratosis | 0.96 | 0.84 | 0.97 | 0.68 | 0.75 |
|  | Basal cell carcinoma | 0.94 | 0.91 | 0.94 | 0.80 | 0.85 |
|  | Melanoma+ | 0.98 | 0.94 | 0.98 | 0.82 | 0.88 |
|  | Surgically eligible melanocytic lesions^ | 0.96 | 0.03 | 0.99 | 0.14 | 0.04 |
|  | Squamous cell carcinoma | 0.98 | 0.85 | 0.98 | 0.80 | 0.82 |
|  | Squamous cell carcinoma in-situ | 0.96 | 0.58 | 0.99 | 0.73 | 0.65 |
|  | Other (benign) | 0.94 | 0.67 | 0.98 | 0.80 | 0.73 |
|  | Other (malignant) | 0.99 | 0.50 | 0.99 | 0.40 | 0.44 |
|  | Other (non-neoplastic, inflammatory, infectious) | 0.98 | 0.29 | 0.99 | 0.63 | 0.40 |

**Supplementary Table 2.**
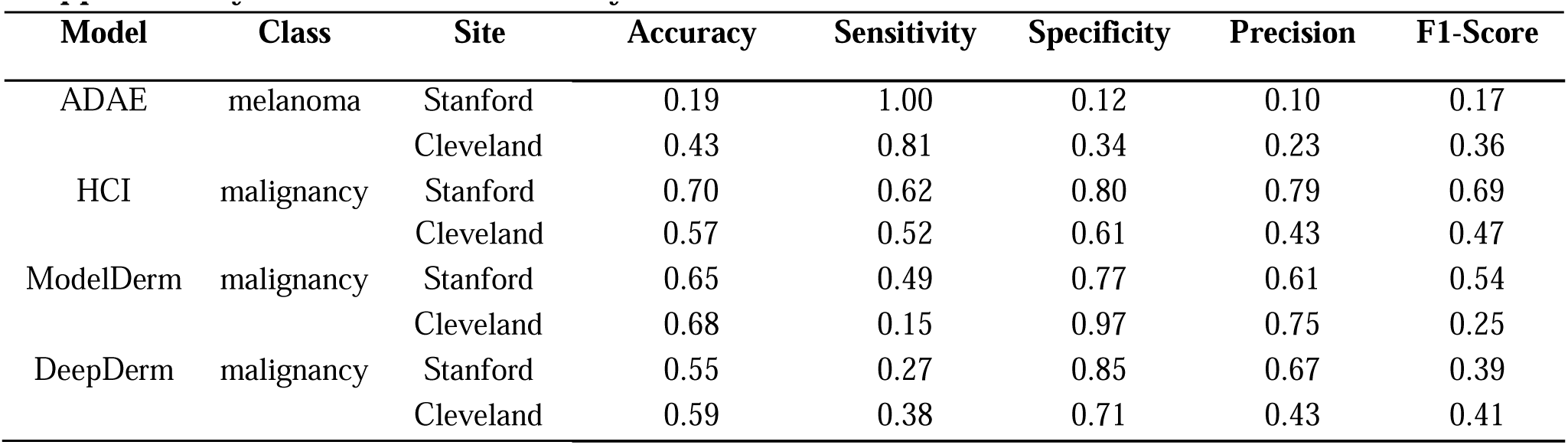
Model subanalysis-Stanford vs Cleveland Clinic.

| Model | Class | Site | Accuracy | Sensitivity | Specificity | Precision | F1-Score |
| --- | --- | --- | --- | --- | --- | --- | --- |
| ADAE | melanoma | Stanford | 0.19 | 1.00 | 0.12 | 0.10 | 0.17 |
|  |  | Cleveland | 0.43 | 0.81 | 0.34 | 0.23 | 0.36 |
| HCI | malignancy | Stanford | 0.70 | 0.62 | 0.80 | 0.79 | 0.69 |
|  |  | Cleveland | 0.57 | 0.52 | 0.61 | 0.43 | 0.47 |
| ModelDerm | malignancy | Stanford | 0.65 | 0.49 | 0.77 | 0.61 | 0.54 |
|  |  | Cleveland | 0.68 | 0.15 | 0.97 | 0.75 | 0.25 |
| DeepDerm | malignancy | Stanford | 0.55 | 0.27 | 0.85 | 0.67 | 0.39 |
|  |  | Cleveland | 0.59 | 0.38 | 0.71 | 0.43 | 0.41 |

**Supplementary Table 3.** Model subanalysis: Demographic Subgroups. Large lesions were defined as length and width of the lesions are above the cut-off value of 6mm. Small lesions are all the other lesions.

| Model | Subgroup | Accuracy | Sensitivity | Specificity | Precision | F1-Score |
| --- | --- | --- | --- | --- | --- | --- |
| ADAE | Melanoma History | 0.28 | 0.93 | 0.12 | 0.21 | 0.34 |
|  | No Melanoma History | 0.19 | 1.00 | 0.13 | 0.07 | 0.13 |
|  | Female | 0.22 | 0.97 | 0.14 | 0.10 | 0.18 |
|  | Male | 0.20 | 0.97 | 0.13 | 0.10 | 0.18 |
| | Age $\geq 66$ | 0.18 | 0.98 | 0.10 | 0.10 | 0.18 |
| | Age $< 66$ | 0.24 | 0.96 | 0.17 | 0.10 | 0.18 |
|  | Fitz 1-2 | 0.21 | 0.97 | 0.13 | 0.11 | 0.19 |
|  | Fitz 3-6 | 0.18 | 1.00 | 0.14 | 0.06 | 0.11 |
|  | White | 0.19 | 1.00 | 0.11 | 0.09 | 0.17 |
|  | Non-White | 0.21 | 0.97 | 0.13 | 0.10 | 0.18 |
| | Small Lesion ($<6\text{mm}$) | 0.18 | 1.00 | 0.12 | 0.08 | 0.14 |
| | Large Lesion ($\geq 6\text{mm}$) | 0.20 | 1.00 | 0.10 | 0.11 | 0.20 |
| ModelDerm | Melanoma History | 0.62 | 0.38 | 0.86 | 0.73 | 0.50 |
|  | No Melanoma History | 0.61 | 0.44 | 0.78 | 0.69 | 0.54 |
|  | Female | 0.62 | 0.37 | 0.84 | 0.66 | 0.48 |
|  | Male | 0.60 | 0.45 | 0.78 | 0.71 | 0.55 |
| | Age $\geq 66$ | 0.58 | 0.48 | 0.73 | 0.75 | 0.59 |
| | Age $< 66$ | 0.64 | 0.33 | 0.85 | 0.59 | 0.43 |
|  | Fitz 1-2 | 0.59 | 0.42 | 0.79 | 0.71 | 0.53 |
|  | Fitz 3-6 | 0.78 | 0.56 | 0.86 | 0.57 | 0.57 |
|  | White | 0.59 | 0.42 | 0.80 | 0.71 | 0.53 |
|  | Non-White | 0.73 | 0.55 | 0.81 | 0.57 | 0.56 |
| | Small Lesion ($<6\text{mm}$) | 0.63 | 0.35 | 0.79 | 0.48 | 0.40 |
| | Large Lesion ($\geq 6\text{mm}$) | 0.69 | 0.64 | 0.74 | 0.73 | 0.68 |
| DeepDerm | Melanoma History | 0.56 | 0.31 | 0.82 | 0.62 | 0.41 |
|  | No Melanoma History | 0.55 | 0.27 | 0.84 | 0.65 | 0.38 |
|  | Female | 0.58 | 0.24 | 0.85 | 0.57 | 0.34 |
|  | Male | 0.54 | 0.29 | 0.83 | 0.67 | 0.40 |
| | Age $\geq 66$ | 0.49 | 0.32 | 0.79 | 0.72 | 0.45 |
| | Age $< 66$ | 0.62 | 0.20 | 0.87 | 0.49 | 0.28 |
|  | Fitz 1-2 | 0.53 | 0.27 | 0.83 | 0.64 | 0.38 |
|  | Fitz 3-6 | 0.77 | 0.44 | 0.90 | 0.62 | 0.51 |
|  | White | 0.52 | 0.26 | 0.84 | 0.67 | 0.38 |
|  | Non-White | 0.68 | 0.38 | 0.83 | 0.53 | 0.44 |
| | Small Lesion ($<6\text{mm}$) | 0.58 | 0.23 | 0.86 | 0.58 | 0.33 |
| | Large Lesion ($\geq 6\text{mm}$) | 0.52 | 0.31 | 0.84 | 0.76 | 0.44 |

## Data Availability

Data produced are available online at https://stanfordaimi.azurewebsites.net/datasets/f4c2020f-801a-42dd-a477-a1a8357ef2a5

